# Developing a video expert panel as a reference standard to evaluate respiratory rate counting in paediatric pneumonia diagnosis: protocol for a cross-sectional study

**DOI:** 10.1101/2022.08.08.22278556

**Authors:** Ahad Mahmud Khan, Salahuddin Ahmed, Nabidul Haque Chowdhury, Md Shafiqul Islam, Eric D McCollum, Carina King, Ting Shi, Kamrun Nahar, Robynne Simpson, Ayaz Ahmed, Md Mozibur Rahman, Abdullah H Baqui, Steven Cunningham, Harry Campbell, RESPIRE Collaboration

**Author notes:** **Corresponding Author:** Ahad Mahmud Khan, The University of Edinburgh, Edinburgh, UK.

## Abstract

**Introduction:** Manual counting of respiratory rate (RR) in children is challenging for health workers and can result in misdiagnosis of pneumonia. Some novel RR counting devices automate the counting of RR and classification of fast breathing. The absence of an appropriate reference standard to evaluate the performance of these devices is a challenge. If good quality videos could be captured, with RR interpretation from these videos systematically conducted by an expert panel, it could act as a reference standard. This study is designed to develop a video expert panel (VEP) as a reference standard to evaluate RR counting for identifying pneumonia in children.

**Methods and analysis:** Using a cross-sectional design, we will enrol children aged 0-59 months presenting with suspected pneumonia at different levels of health facilities in Dhaka and Sylhet, Bangladesh. We will video record a physician/health worker counting RR manually and also using an automated RR counter (ChARM) from each child. We will establish a standard operating procedure for capturing quality videos, make a set of reference videos, and train and standardise the VEP members using the reference videos. After that, we will assess the performance of the VEP as a reference standard to evaluate RR counting. We will calculate the mean difference and proportions of agreement within ±2 breaths per minute and create Bland-Altman plots with limits of agreement between VEP members.

**Ethics and dissemination:** The study protocol was approved by the National Research Ethics Committee of Bangladesh Medical Research Council (BMRC), Bangladesh (registration number: 39315022021) and Edinburgh Medical School Research Ethics Committee (EMREC), Edinburgh, UK (REC Reference: 21-EMREC-040). Dissemination of the study findings will be through conference presentations and publications in peer-reviewed scientific journals.

**Strengths and limitations of this study:**

- Expert paediatricians will provide feedback to develop a standard operating procedure for videography of child chest movements.
- The video expert panel will be trained and standardized using the expert paediatrician-interpreted reference videos.
- Video expert panel members will be masked to respiratory rate counted by each other and to respiratory rate manual counts, and with the automated counter.
- Children with varying severity of illness will be enrolled from different levels of health facilities in Bangladesh.
- Despite the availability of multiple respiratory rate counters, only the ChARM device will be used in this study.

## Introduction

Pneumonia is one of the leading causes of mortality in children aged below five years worldwide.^1^ Approximately 68 million pneumonia episodes and 0.65 million deaths occur annually in under-five children due to pneumonia.^2^ The highest number of deaths due to pneumonia among children below five years were in sub-Saharan Africa, South Asia, and Southeast Asia. There were an estimated 2.7 million episodes of pneumonia in Bangladesh and 21,275 deaths due to childhood pneumonia in 2015.^3^

According to the World Health Organization (WHO) guidelines, pneumonia diagnosis in children for frontline health workers is primarily based on increased respiratory rate (RR) and/or chest indrawing.^4 5^ Fast breathing is the most common sign of pneumonia. It is most commonly identified by counting the RR manually.^5^ However, manual counting can be challenging for the health workers^6^, often leading to incorrect diagnosis and, consequently, inappropriate treatment.^7-9^

Better performing RR diagnostics to support frontline health workers might improve the diagnosis of pneumonia. Three automated RR counting devices are considered suitable for use by health workers, i.e., Children ‘s Automated Respiration Monitor (ChARM)^10 11^, Rad-G^12 13^ and uPM60^14^. The Philips ChARM converts chest movements detected by accelerometers into a precise breathing count using specially designed algorithms. The device is placed around the child ‘s belly, and it automatically counts RR and classifies fast breathing according to WHO guidelines^10 11^. The usability and acceptability of this device have been tested in a study in Ethiopia^15^ and Nepal.^16^

A reference standard is essential to evaluate the performance of RR counters in field settings. However, the absence of an appropriate reference standard is a challenge. Different reference standards have been used in various studies.^17 18^ Most of the existing studies have used manual RR count by an expert person (e.g., physician, nurse) as the reference standard.^19^ Few studies have been found using automated monitors.^20 21^ The possible biases using a human expert ‘s count as the reference standard include difficulty measuring the RR over the same simultaneous period and inconsistencies in human expert RR counting.^6^ The automated monitors do not measure chest movements directly, but other variables such as carbon dioxide (CO2), pulse oximeter signal or photoplethysmogram (PPG), sound etc., are used to extract RR.^19 22-24^

Videography of a child ‘s chest movements and interpretation by the experts could be used as a reference standard.^17 18 25^ If quality videos could be recorded and the interpretation of RR from these videos could be systematically conducted by a video expert panel (VEP), it could be an ideal and non-biased reference standard. This study aims to develop a VEP as a reference standard for evaluating RR counting manually and using ChARM in paediatric pneumonia diagnosis.

## Study objectives

### Primary objectives

1. To develop a standard operating procedure (SOP) for video recording a child ‘s chest movements for RR interpretation.
2. To create a set of reference videos for the training and standardisation of the VEP members.
3. To train and standardise the VEP members to interpret RR using the reference videos.
4. To assess the performance of the VEP to evaluate RR counting manually and with ChARM based on videos captured in real-world settings.

### Secondary objectives

1. To assess the accuracy of the ChARM device in counting RR compared to the VEP as the reference.
2. To determine the duration of counting RR using the ChARM device.
3. To explore the potential influence of the ChARM device on RR count compared to standard observation techniques.

## Methods and analysis

### Study design and settings

This will be a cross-sectional study. First, we will capture videos of child chest movements, develop a SOP of videography, make a set of reference videos and use these reference videos to train and standardize the VEP. We will record these videos from the inpatient department (IPD) of the Institute of Child and Mother Health (ICMH), Dhaka, Bangladesh. After that, we will use the VEP as a reference standard to evaluate RR counting. The videos will be recorded from different levels of health facilities in Bangladesh. Three community clinics (CCs) and a sub-district hospital (Zakiganj Upazila Health Complex) in Sylhet and ICMH in Dhaka will be selected. CCs are the lower-level health facility in Bangladesh staffed by community healthcare providers (CHCP).^26^ Patients from the CCs are referred to sub-district hospitals. Patients often go to sub-district hospitals directly.^27^

### Study population

#### Inclusion criteria

i. Age <2 months presenting with any illness
ii. Age 2-59 months presenting with cough and/or difficulty breathing

#### Exclusion criteria

i. Presence of any danger sign (unable to drink or feed, vomit everything, convulsions, lethargic or unconscious)
ii. Unwilling to provide written informed consent

### Study procedures

#### Enrolment and consenting

In the hospital, a physician will screen all children who will visit the outpatient clinic (sub-district hospital) or are already admitted to the inpatient department (ICMH) and enrol eligible children. In the CC, the CHCP will screen every child who will visit his/her CC. If a child becomes eligible, the CHCP will enrol the child and contact research staff who will visit the CC immediately and will complete data collection. Informed written consent will be obtained from the child ‘s mother, father, or other available caregiver before data collection.

#### Developing SOP of videography

Research staff will record videos of child chest movements from 30 children. The Canon EOS M50 camera^28^ will be used for recording videos. If a good source of natural light is not available, we will use a light-emitting diode (LED) lamp for lighting. The child ‘s clothes will be removed from the lower end of the neck through the umbilicus so that the chest and belly are exposed. The child ‘s face or any other identifiable part will not be recorded. The videos will be recorded from different camera positions and angles with varying levels of light. The shots will be taken either using a tripod or not using it. Video recording will be done both when counting RR manually twice and using the ChARM device twice.

Once the child is calm, the research staff will start video recording, and the physician will tap the microphone with his/her finger and begin counting RR. The physician will count RR for one minute and tap the microphone again when counting is finished, and the research staff will stop the recording. After that, the physician will attach the ChARM device, and the research staff will start video recording. Once the child is calm, the physician will start the ChARM device and also begin counting RR manually, and the video recording will be done in parallel to the physician ‘s use of ChARM like before. The physician will count the total number of breaths until ChARM completes its counting. The ChARM device will show a green or red signal when it finishes its count. The video recording will continue until the device is done. The time taken by the ChARM device to get the RR will be recorded. A maximum of 3 minutes will be allowed for the ChARM to register a reading or display an error message. Otherwise, it will be documented as a failed attempt.

Each video file will be saved in the camera with a file name. The research staff will note the file name in the data collection sheet. Then the videos will be downloaded from the camera and saved on a password-protected computer. After that he will edit the video by removing additional portions of the recording, removing sound or identifiable features, and reducing file size. Video Editing Software (Adobe Premiere Pro) will be used for video editing.

There will be two expert paediatricians (i.e., medical practitioners having a post-graduate degree in paediatrics and specialized in treating sick children). These videos will be sent to them for their feedback on video quality (decision of using tripod, camera positions, angles, light, or other issues) and to count RR. Their feedback will be incorporated, and the video recording procedures will be modified accordingly. A SOP for videography will be developed to maintain consistency across all study hospitals/clinics.

Figure 1 shows the schematic diagram of developing the SOP for videography.

**Figure 1:**
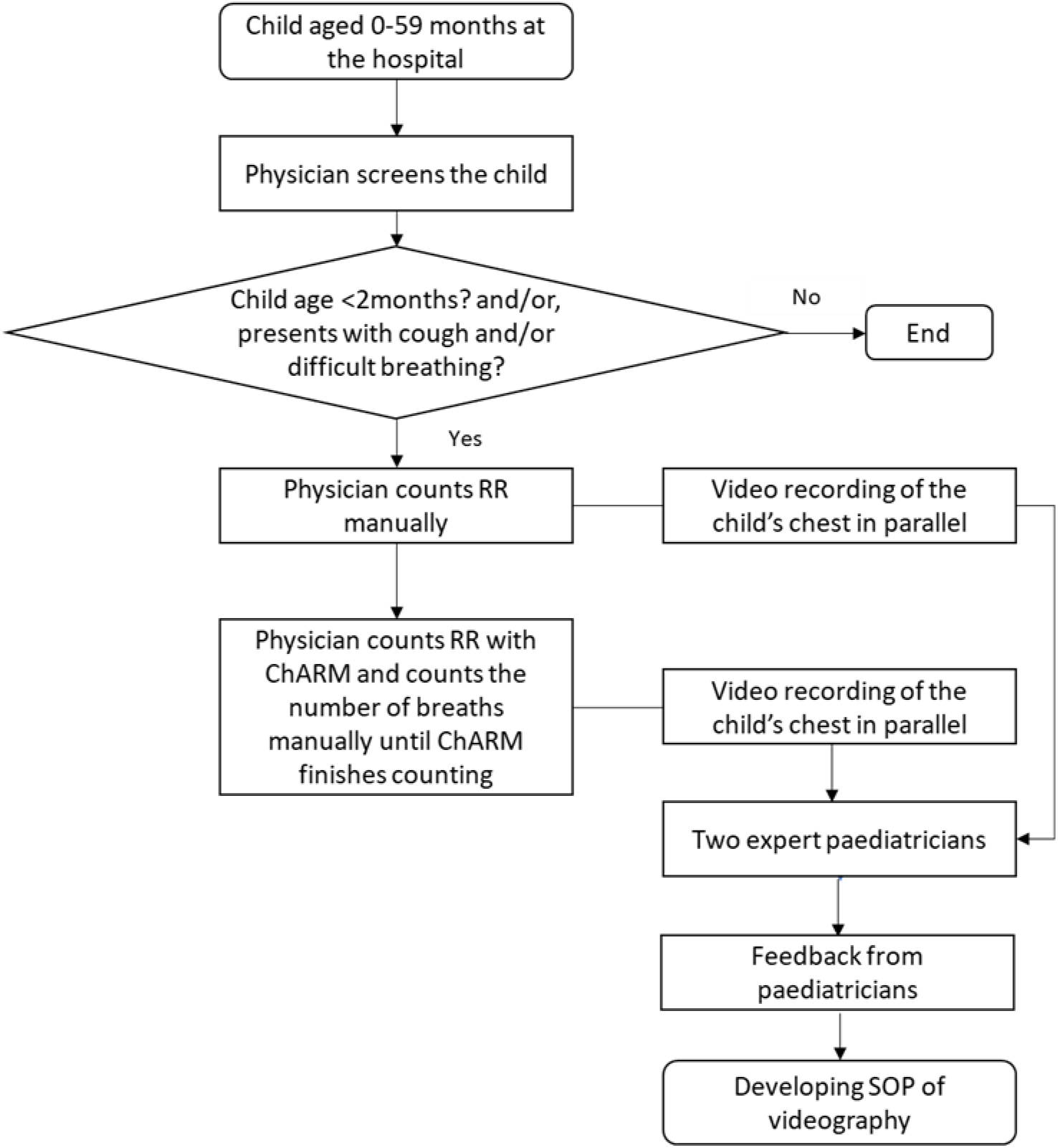
Developing SOP of videography

#### Making a set of reference videos

Following the development of the SOP, videos will be recorded from 50 children using this standardised procedure. A physician will count the RR manually once and with the ChARM device once, and video recording will be done simultaneously. The same procedure described in Figure 1 will be followed. If the physician fails in their first attempt to have a satisfactory recording, they will make two more attempts for each manual and ChARM counts. The reason for the attempted failure will be documented. The condition of the child during RR measurement will also be noted.

There will be three expert paediatricians. Each video will randomly be allocated to two of the paediatricians. If both paediatricians disagree, i.e., disagreement in readability or difference of RR >±2 breaths per minute (bpm), then the video will be sent to the third paediatrician. If any two paediatricians agree, i.e., agreement in interpretability and difference of RR ≤±2 bpm, the videos will be considered reference videos. The mean RR count of paediatricians in agreement will be considered as the final RR count.

Figure 2 presents the schematic diagram of making a set of reference videos.

**Figure 2:**
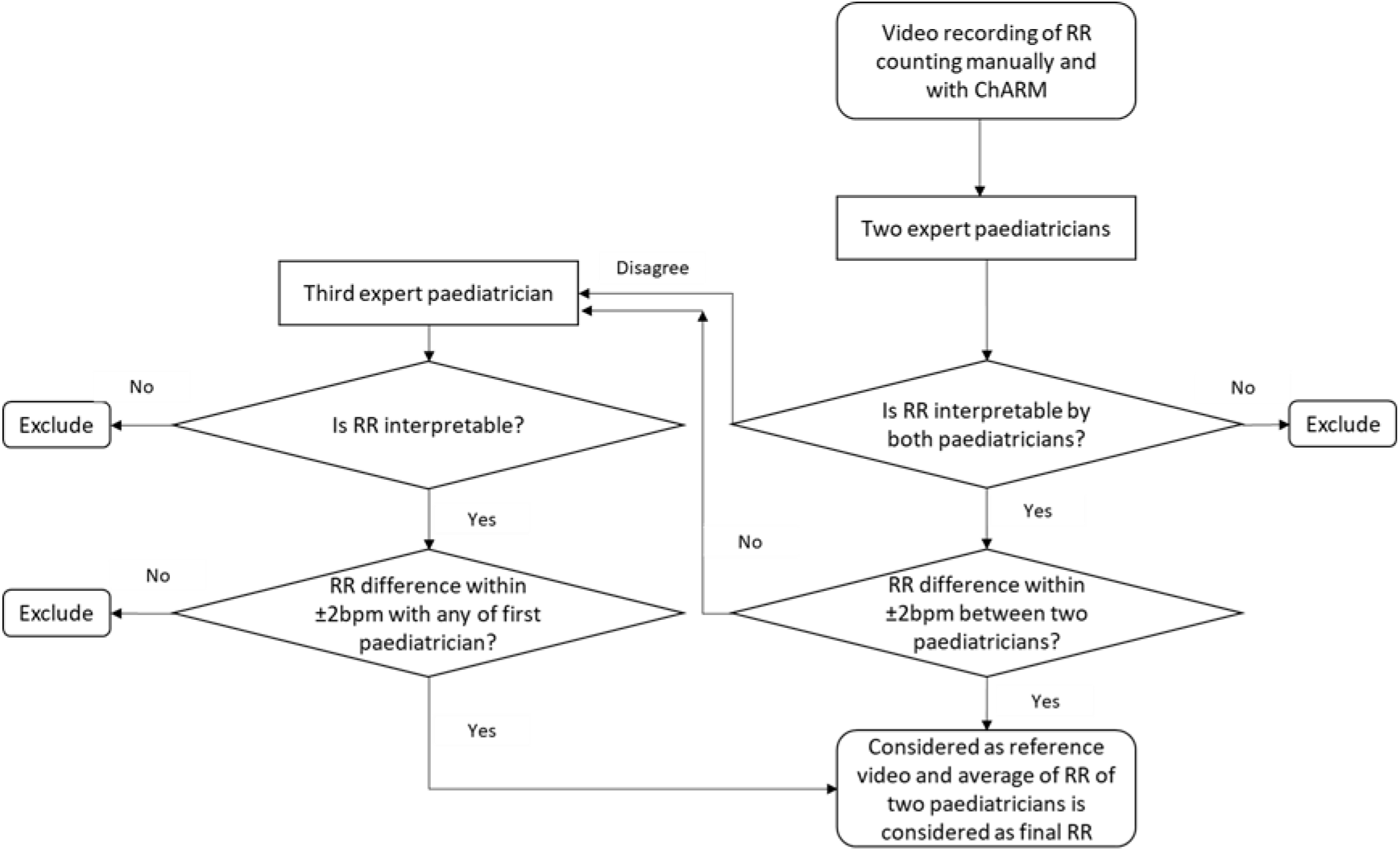
Making a set of reference videos

#### Training and standardisation of VEP members

Six local physicians with a MBBS degree will be invited for training and standardisation. The physicians will be trained on counting RR using videos of known RR counts from reference videos. The Principal Investigator (PI) will conduct the training sessions. After completion of training, they will be evaluated individually in a blinded manner. Twenty videos will be assigned to each VEP member from the reference videos. Those physicians who achieve a pass mark (at least 80%) in the evaluation will be considered to work as VEP members in the study.

#### Evaluating the performance of VEP

The performance of VEP will be evaluated using the videos recorded in different levels of health facilities (CCs, sub-district hospital and ICMH). RR of each child will be assessed twice, i.e., manually and using ChARM by the CHCP in the CC and by a physician in the hospital (as described in Figure 1). The videos will be recorded following the SOP. Each video will be randomly assigned to two VEP members to interpret the RR. If there is an agreement of being uninterpretable, the videos will be excluded. If the RR is ≤±2 bpm between both members, their average RR will be considered final. However, if there is a disagreement in interpretability or the difference of the RR count is >2 bpm between the first two members, the video will be sent to a third-panel member. If there are inconsistencies among all three members (if the RR is not ≤2 bpm between any two members), the videos will be sent to a fourth-panel member. If there is agreement between the fourth-panel member and any other member who read the video earlier, their average RR will be considered final. However, if there is no agreement between any two panel members, the videos will be excluded from the analysis (Figure 3).

**Figure 3:**
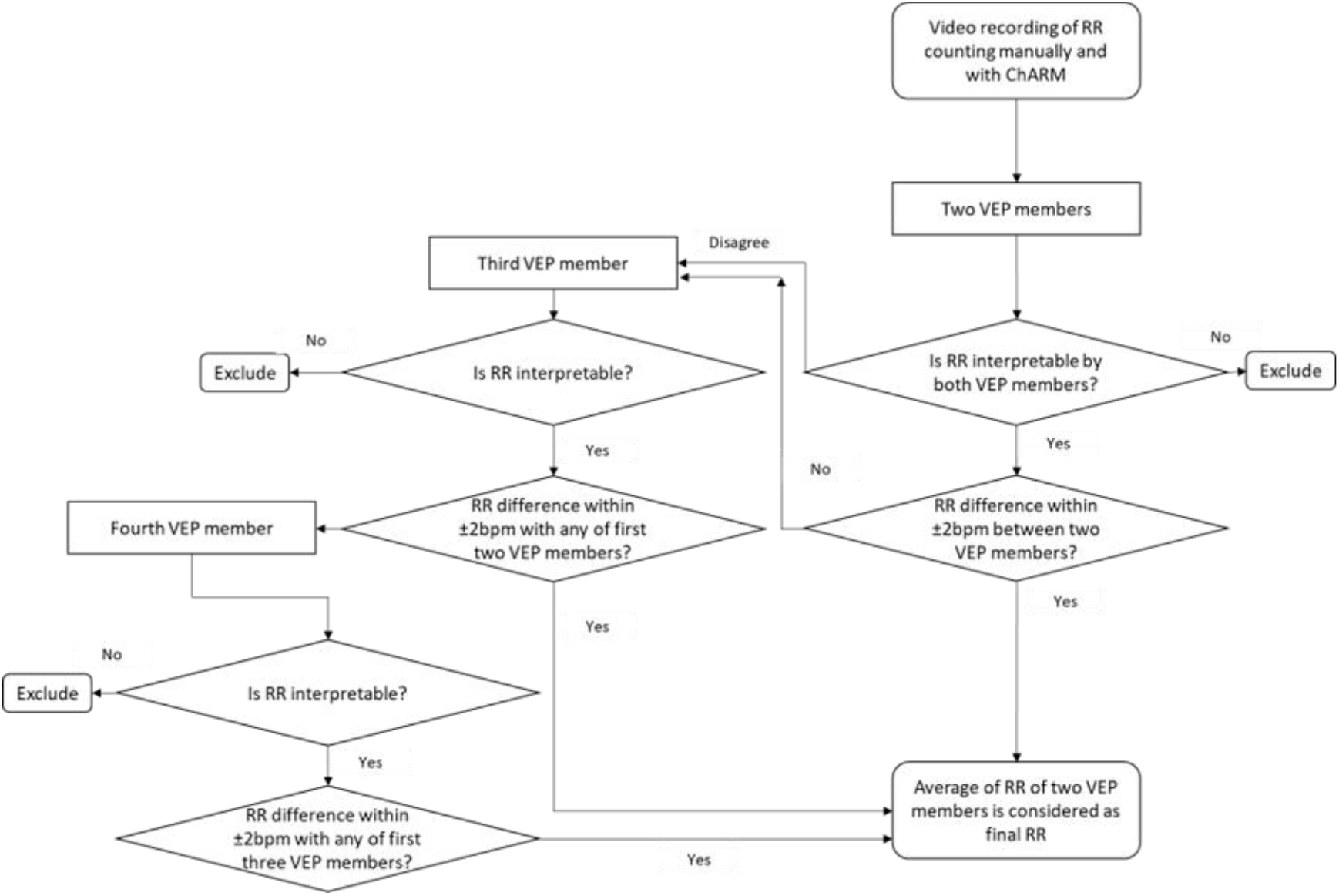
Respiratory rate interpretation from video recording by video expert panel

#### Quality control measures

The videos will be checked periodically to ensure quality by the PI. Any quality issues will be reported, and corrective feedback will be provided. The expert paediatricians will view a 10% random sample of videos in which they are blinded to the panel ‘s final classification. The panel ‘s overall performance will be evaluated using the expert ‘s interpretation as the reference. Lastly, to monitor individual member interpretation reproducibility, all members will randomly be reassigned 20% of images they interpreted previously to check intra-reader agreement.

#### Development of a web-based automated system

A web-based automated system will be developed. This system will use the Hypertext Preprocessor (PHP) platform for the user interface and the Structured Query Language (SQL) platform for the database. The purpose of developing this system is an automated distribution of the video recordings among the expert paediatricians and VEP members, the input of readings by the members and the generation of automated reports. The system will be password-protected and connected to a secured server. The expert paediatricians and VEP members will use this program. Each will need to log in with their user ID and password. Access to each expert paediatrician and VEP member will be limited to what they are assigned. When logged in, they will see the list of recordings assigned to them. They will open the video on the computers and interpret the RR. The VEP members will be blinded to other members ‘ readings.

### Operational definitions

#### Interpretability

The video will be considered ‘interpretable ‘ when the expert paediatrician or VEP member will be able to view the child ‘s chest movements and measure RR for the whole duration. On the other hand, a video will be counted as ‘uninterpretable ‘ if the physician will not be able to view and count RR for the whole duration of the recording. The videos can be unreadable for various reasons, e.g., wrong position and angle of the camera, inappropriate focus, poor lighting, inadequate exposure or if the child is agitated, moving, crying etc.

#### Agreement and disagreement

The agreement will be defined when two expert paediatricians or VEP members interpret the video as

- uninterpretable or
- interpretable and the difference between RR count ≤±2 bpm.

Disagreement is when

- one expert paediatrician/VEP member interprets the video as interpretable, and another interprets it as unreadable or
- the difference between their RR count is >2 bpm.

### Data collection and storage

Field data will be collected in paper forms. After entry, data will be transferred to a password-protected server (SQL Server 2008 R2) located in Dhaka, Bangladesh, in real-time using internet connectivity. Recorded videos will be transferred to encrypted OneDrive/Dropbox cloud storage. The VEP members will interpret RR from recorded videos using the web-based automated software, and the data will also be stored on the server in real-time. De-identified data will be transferred and stored on the DataShare server at the University of Edinburgh, UK. Data collection started on 6 December 2021 and is planned to be completed by October 2022.

### Sample size calculation

The videos from the first 30 children will be used to establish the best procedure for capturing videos. After that, the videos from 50 children will be used for making a set of reference videos to train and standardize VEP. To evaluate the performance of VEP, Bland-Altman ‘s statistical methods for assessing agreement between two methods of clinical measurement are used.^29 30^ A total of 226 interpretable videos of each method (manual and ChARM) are required, which will provide 90% power to assess agreement in RR counts between two VEP members, considering type-I error α=0.05, expected mean difference ±0.5, expected SD 1.5 and maximum allowed difference 4. Assuming 80% interpretability of the videos and 20% failure to record videos, the required number of children is about 350.

### Statistical analysis

To standardize the VEP members, we will estimate the mean difference of RR counts and the percent agreement within ±2bpm between each VEP member and expert paediatricians. We will consider expert paediatricians ‘ counts as the gold standard. To assess the performance of VEP members, we will produce Bland-Altman plots^29^ to assess the agreement in RR counts and calculate the percent agreement between primary panel members. We will also assess the intra-reader agreement of each VEP member. We will use Cohen ‘s kappa statistic to estimate the inter-reader agreement of identifying fast breathing between panel members. We will also measure the mean difference of RR counts and the percent agreement within ±2bpm between each VEP member and expert paediatricians from QC data.

## Data Availability

All data produced will be available online at the DataStore repository at the University of Edinburgh, UK.

## Ethics and dissemination

Ethical approval was obtained from the National Research Ethics Committee of Bangladesh Medical Research Council (BMRC), Bangladesh (Registration Number: 39315022021), and Edinburgh Medical School Research Ethics Committee (EMREC), Edinburgh, UK (REC Reference: 21-EMREC-040). Informed written consent will be taken from the parent or guardian of each child. Dissemination will be through conference presentations and publications in peer-reviewed journals. Anonymised data files will also be stored securely in the DataStore repository at the University of Edinburgh, UK and will be shared after the publication the main paper.

## Acknowledgments

The authors are grateful for the support and contributions of Dr Arunangshu Dutta Roy, Asim Nehal, Rizouan Ur Rashid, Rakib Bhuiyan, Dr Nusrat Sharmin Asma, Dr Farjana Hossain, Dr Muhammad Shariful Islam, Dr Md. Jahurul Islam, and the Ministry of Health and Family Welfare, Government of Bangladesh.

## Footnotes

### Author Contributions

AMK and HC conceptualised and designed this study. HC, SC, AHB, EDM and TS provided mentorship to AMK. HC, SC and EDM critically reviewed the study design. KN, RMS and AA will review the videos and provide feedback to video quality. NHC and MSI will be responsible for data management. AMK drafted the manuscript, and all authors critically reviewed and approved the final manuscript before submission. The RESPIRE collaboration comprises the UK Grant holders, Partners and research teams as listed on the RESPIRE website (www.ed.ac.uk/usher/respire) including Linda Bauld.

### Patient and public involvement

Patients and/or the public were not involved in this research ‘s design, conduct, reporting, or dissemination plans.

